# WASTE DISPOSAL MECHANISM AS DETERMINANT OF HEALTH STATUS OF RESIDENTS OF IJEBU-ODE METROPOLIS

**DOI:** 10.1101/2025.08.21.25332252

**Authors:** James Sunday Ajayi, Zacchaeus Oluseyi Ojekunle

**Affiliations:** Department of Geography and Environment Management, Tai Solarin University of Education Ijagun; Department of Environmental Management and Toxicology, Federal University of Agriculture, Abeokuta

**Keywords:** Waste management, mechanism, health related diseases

## Abstract

Descriptive survey design was adopted for the study. One hundred and twenty (n=120) participants were drawn using simple random sampling technique from ten streets of Ijebu Ode metropolis. Interview and personal observation were used to collect the data. These data collections were Residents’ Waste Management Methods, Solid Waste Management Attitude Scale, Key Informant Interview Schedule. A mean score of 2.0 and above indicates that the residents are of positive attitude towards solid waste management while a mean score of less than 2.0 indicates negative attitude. The use of single bin in collection of waste appears inadequate coupled with twice frequency of disposal of wastes in a month. Given the result (median = 2.00) wastes from homes of the residents were disposed on a generally note twice a month, and a score (median 1.00) indicate that common waste receptacles placed in designated locations are the only procedure of waste disposal among the residents. On awareness of health related diseases, the results (Median= 1.00) residents of Ijebu-Ode community claimed awareness of health related diseases of waste disposal and result of (Median = 1.00) shoes that the frequency of outbreak of diseases in keeping refuse dumps among the residents was low.

It was concluded that the residents claimed of awareness of waste management and health related diseases but it obvious that the residents do not have full understanding or knowledge of waste management but rather base on residual knowledge and past experience on waste management which is still accounting for finding the environment be littered and polluted with wastes. Among others, there should be public education on proper ways of solid waste disposal in the metropolis to inform the general public on the implications of unhealthy environment and the need to keep their communities clean. The education could be done by Local Authorities and the Ogun State Environmental Programme Agency (OGEPA) was recommended.

## INTRODUCTION

Health is the level of functional and metabolic efficiency of a living organism. According to Marmot, (2007) health is a universal human aspiration and a basic human need. The development of society, rich or poor, can be judged by the quality of its population’s health, how fairly health is distributed across the social spectrum, and the degree of protection provided from disadvantage due to ill-health (Marmot, 2007). The World Health Organization (WHO) as cited in Grad, (2002) defined human health as a state of complete physical, mental, and social well-being and not merely the absence of disease or infirmity. Generally, the context in which an individual lives is of great importance for both his health status and quality of their life (Simandan, 2018). It is increasingly recognized that health is maintained and improved not only through the advancement and application of health science, but also through the efforts and intelligent lifestyle choices of the individual and society. According to the World Health Organization (WHO, 2011), the main determinants of health include the social and economic environment, the physical environment, and the person’s individual characteristics and behaviors.

More specifically, the key factors that have been found to whether determine or not people are healthy or unhealthy include social and physical environment, personal health practices and coping skills, healthy child development, biology and genetics, health care services, gender and culture. However, the environment is often cited as an important factor influencing the health status of individuals. This involves characteristics of the natural environment, the built environment, and the social environment. Hygiene factors such as clean water and air, adequate housing, and safe communities and roads all have been found to contribute to good health.

Hygiene is a necessary condition for an individual to be healthy. A healthy person is one that can be economically productive and valuable. Hygiene implies cleanliness. The significance of hygiene is embedded in a common saying that cleanliness is next to Godliness. Environmental hygiene is about the measures taken to keep the human environment safe and healthy and these include waste disposal, clean water supplies, food safety controls and good housing. Hygiene implies a set of practices performed to preserve health. According to the World Health Organization (WHO), “Hygiene refers to conditions and practices that help to maintain health and prevent the spread of diseases. An hygienic environment is the environment that is safe from all environmental threats such as pollution by solid, liquid and gaseous wastes that may cause different pathogenic infections such as malaria, cholera and so on that may affect human.

Theoretically, hygienic environment is needed for a safety and conducive environment. Hygienic environment is free from any problem or threat, encourages healthy living environment and other environmental disaster cannot be envisaged. Waste as the term implies, is any solid, liquid or gaseous substance or material which being a scrap or being super flows, refuse or reject, disposed-off or required to dispose as unwanted. In countries around the world, one major environmental problem that confronts municipal authorities is solid waste disposal. Most government authorities are confronted by mounting problems regarding the collection and disposal of solid wastes. Today, municipal solid waste collection and disposal are particularly problematic in developing country cities, but many Western cities have also grappled with this problem in the past and some probably still do (Boniface, 2013).

Waste is more easily recognized than defined (Banjo, Adebambo & Dairo, 2009). Material objects can become waste when it is no longer useful to the owner or it is used and fails to fulfill its purpose (Gourlay, 1993). A great mixture of substances including fine dust, cinder, metal, glass, paper and cardboard, textiles, putrescible vegetable materials and plastic characterise solid waste (Simmens, 1981 as cited in Banjo, et. al., 2009). As time passes accumulation of waste outstrips its control. There is no single solution to the challenge of waste management, process is usually framed in term of generation, storage, treatment and disposal, with transportation inserted between stages of required. Hence, a combination of source reduction, recycling, incineration and burying in landfills and conversion is currently the optimal way to manage domestic wastes (George, 2008).

In Africa, the generation of waste and its disposal; both domestic and industrial, continues to increase in cycle with growth in consumption and its association with health problems (Achankeng, 2003). For instance, per capita waste generation increased nearly three-folds over the last two decades on the continent, reaching a higher level than that in developed countries (UNEP, 2009). However, out of these huge waste generations on the continent of Africa, countries spend as much as 20 to 50 percent of their district, municipal or metropolitan revenues on waste management. Yet, two-thirds of the solid waste generated is not collected (Da-Zhu, Asnani, Zurbugg, Anapolsky & Mani, 2008). Such inadequate waste collection and disposal practices create problems on the environment as well as human health and consequently economic and other welfare losses. Even the collected waste ends up in an uncontrolled dump sites or burnt (Achankeng, 2003, cited in Abagale, Mensah & Agyeman-Osei, 2012).

United States, Environmental Protection Agency (2009) and World Health Organisation (2009) argue that the major cause of increase in solid waste has been the advent of modernization, technological advancement and increase in global population. These have created rise in demand for food and other human needs, which has also culminated into increase in the amount of waste being generated daily by individual households. Improper solid waste disposal contributes to environmental health problems in both developed and developing countries. This is because the health implications of improper solid waste disposal can be detrimental to people exposed to such situations especially school children, waste workers, and workers in facilities producing toxic and contagious material. Other high-risk groups include population living near a waste dump and those, whose water supply has become contaminated due to either waste dumping or leakage from landfill sites. Uncollected solid waste also increases risk of injury and infection (Danso-Manu, 2011).

Moreso, according to MLGRD, (2010) as cited in Abul, 2010, it is estimated that in Africa about 70 per cent outpatient cases are sanitation and environmental related diseases. Consequently, improper solid waste disposal have a very high economic and social cost on public health services, as has been estimated by governments, industries, and families.

Navez-Bounchaire (1993) stated that the management of household refuse is tied to perceptions and socio-cultural practices which result in modes of appropriation of space with greatly differenced according to whether the space is private or public. This is relevant to the study because the area has diverse socio-cultural practices, as the population is heterogeneous. Also, Benneh, Songsore, Nabila, Amuzu and Tutu-Yangyuorn (1993) observed that residential domestic waste forms the bulk of all sources of solid waste produced in Urban areas. These household wastes are known to have high densities with high moisture content and the organic component of solid waste, which properly account for about 60% to 80% while tins, cans and paper are probably responsible for about 10% to 20% of the total waste produced.

According to Agbola (1993) cultural derivatives, beliefs, perception and attitudes are learned response sets. They can therefore be modified or changed through education. This points to the facts that people’s unconcerned attitudes toward the waste can be changed for better through education.

However, from past research works it was observed that solid waste is the result of human activities but, serious environmental threat also if not managed properly. The challenge of Solid Waste Management in Ijebu-Ode has always been one of the most pressing challenges. The magnitude of the problem increases with fast urbanization and due to lack of new age technology/machine and inadequate expert/qualified personnel in this field. In Ogun State like other states in Nigeria, the municipalities have established solid waste management system but this system has been extremely outdated, not well managed and supervised properly and inappropriate to meet the demand. Presently none of the municipalities in Ogun State (Ijebu-Ode to be precise) have a technologically-driven recycling and waste disposal system in place.

Also, despite the Local Government effort to clean up the town, the garbage accumulation situation in the town continues to rise day by day. The situation is made even worse by the presence of plastic bottles and polythene papers which are visible in every part of the city. Besides that, piles of garbage are seen on streets and road sides and incidentally constitute an eyesore to visitors coming into the city for the first time. Further, the concentration of population and business activities in the city and the accompanying rapid increase in the volume of solid waste generated from production and consumption activities have led to the prevailing situation. Consequently, the dwellers are facing threat of serious health and environmental problems due to fraternization of waste leachate into ground water and spread of diseases and odour from heaps of waste. Solid waste management has become a global issue and it is now an established fact that waste management is not just a technical issue but it has socio-environmental and cultural dimensions that need solutions through innovative policies, administrative re-organizations, organizational arrangements and community awareness and participation. However, the researcher seeks to study the waste management mechanism adopted by inhabitants of Ijebu-Ode as a determinant of their health status.

The study intends to determine the extent to which waste management mechanism by the inhabitants of *Ijebu*-Ode metropolis predict their health status. The specific objectives of this study are;

1. to ascertain the attitudes of people toward domestic waste management
2. to determine various methods of waste disposal and management are impacted by the levels of education/awareness.
3. determine the adequacy of health status of Ijebu Ode residenc
4. investigate the influence of location of household on disposal of solid waste

## METHODOLOGY

Ijebu-Ode is an ancient city in the south western part of Nigeria, located about latitude 6° 47° and longitude 3° 58°E with an area of 192 km^2^ and a population of 154, 032 at 2006 census, it is bounded in the North by Ijebu North, bounded in the East by Ijebu East Local Government, bounded in the West by Odogbolu Local Government and in the South by Epe Local Government Council of Lagos State. (Aiyelokun and Odekoya 2016).Ijebu-Ode is an ancient city, which is centrally located in relation to other human settlements around it (Bakare, *et. al*., 2011). The city right from the ancient time till date remains the capital of the Ijebu kingdom i.e. the head of all Ijebu speaking towns and villages which are all within the radius of about 20km from Ijebu ode (Ogunowo 2004).

**Figure 1.**
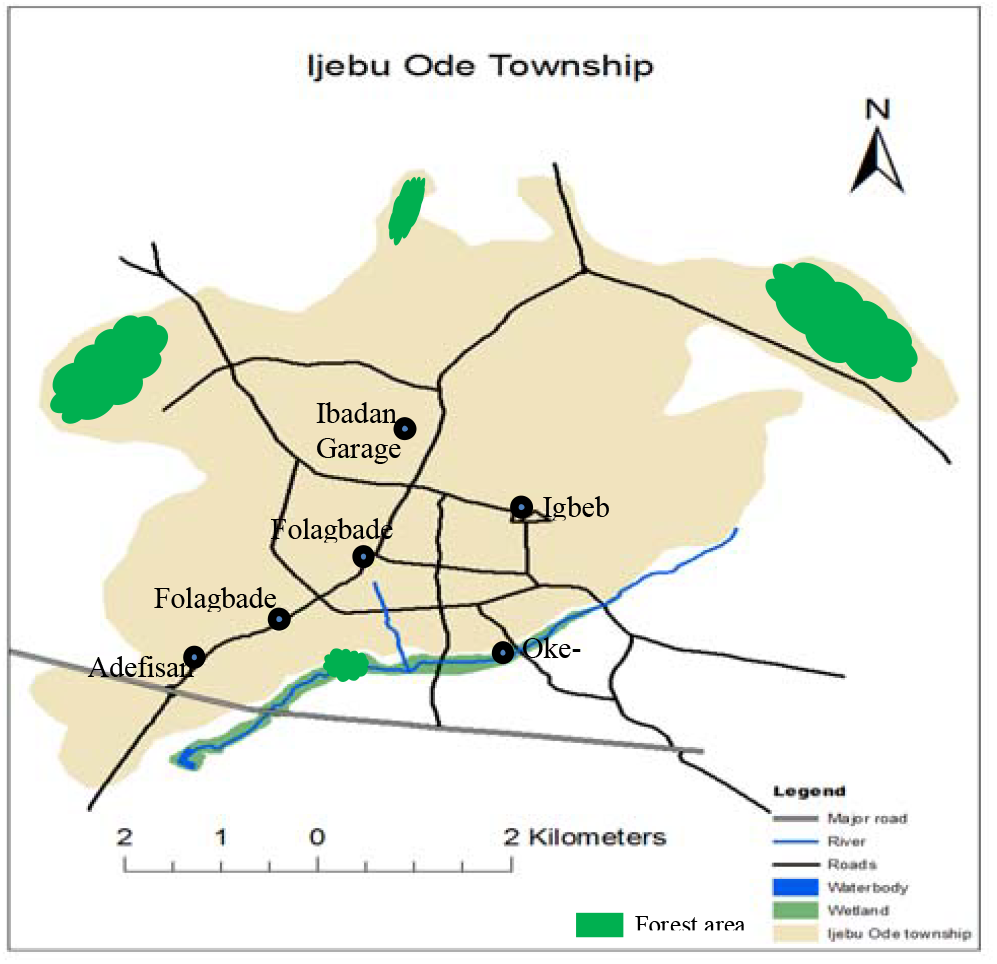
Map of Ijebu ode.

The climate of Ijebu-Ode is within tropical humid climatic zone of Nigeria generally characterize by high rainfall and high relative humidity (Solanke, 2015) Ijebu-Ode’s climatic condition is of alternate wet (April to October) and dry seasons from (November to March). The mean annual rainfall is between 1523 mm and 2340 mm, while the temperature ranges between 250C and 320C with the average annual temperature of about 27^°^C (Bakare,et. al., 2011).

Ijebu-Ode lies squarely within the tropical lowland rain forest region (Mabogunje and Kates 2004). The natural vegetation consists of a great variety of species arranged in a complex verticalstructure with an emergent layer of large trees (up to 60 meters high) including mahogany (*Khayaentandrophragma*), obeche (*Triplochiton*), afara (*Terminalia*), iroko (*Chlorophora*), african walnut (*Lovoatrichilioides*), and ekki (*Lophiraalata*) which form the basis of the major timber industry in the vicinity of the city (Mabogunje and kates 2004).

A sample of one hundred and twenty respondents (n=120) were randomly selected from five (5) wards out of the fifteen (15) in Ijebu-Ode town. Two (2) streets were selected from each ward, making ten streets. The streets are; Porogun I, Itantebo/Ita-Ogbin, Itamapako, Isiwo and Odo-Egba/Oliworo respectively. From each ward, twelve (12) respondents at ten (10) per street were selected. Selections at both levels were by simple random sampling technique. The data collected were analyzed using mean rating, median and standard deviation while the hypothesis formulated in the study were analyzed using analyzed using Spearman’s rho.

## RESULTS AND DISCUSSION

What is the attitude of the residents of Ijebu-Ode community towards solid waste management?

Ground rule: A mean score of 2.0 and above is an indication of positive attitude towards solid waste management while a mean score of less than 2.0 indicates negative attitude.

Looking at table 1, the attitude of residents of Ijebu-Ode community towards solid waste management was positive (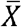 = 2.29). Item by item analysis indicates that the residents play a significant role in the management of waste in their awareness (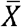= 2.72) just as they claim awareness (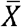= 2.67) of regular collection of garbage health issues.

**Table 1.** Mean and Standard deviation indicating the attitude of residents of Ijebu-Ode community towards solid waste management

| S/N | Items | $\bar{X}$ | S.D | Decision |
| --- | --- | --- | --- | --- |
| 1 | I play an important role in the management of waste in my locality | 2.72 | .60 | Positive |
| 2 | Environmental education should be taught in schools | 2.73 | .67 | Positive |
| 3 | The purchase decisions that I make can increase or decrease the amount of waste my household must get rid of (dispose of) | 2.30 | .87 | Positive |
| 4 | I don't care that burning waste can be bad for my health and the health of others | 2.53 | .82 | Positive |
| 5 | People throw waste on the streets and in the drains and gullies because they have no other means of getting rid of (disposing of) their waste | 1.89 | .95 | Negative |
| 6 | The local Government is not doing enough to fix the waste problem | 1.59 | .68 | Negative |
| 7 | Correct waste management should not be taught in schools | 2.34 | .90 | Positive |
| 8 | Other personal issues (like crime, unemployment and cost of living) are more important to me than a garbage-free community | 2.13 | .95 | Positive |
| 9 | Regular collection of garbage is the only solution to the garbage problem. | 2.67 | .73 | Positive |
| 10 | Picking up waste around my community is my fundamental responsibility as a citizen | 2.04 | .94 | Positive |

Aggregate Mean = 2.29

The result further indicate a positive attitude (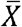=2.73) of residents of Ijebu ode community towards introduction and sustenance of environmental education bring taught in schools. The residents did not agree (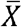=2.53) with indiscriminate burning of solid waste in the open without regards to health of themselves and others. They further demonstrates awareness (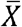= 2.30) for the influence of purchases made to the amount of wastes to dispose from their households.

Further results from table 4.1 indicates positive attitudes (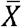= 2.34) of the residents in terms of agreement to the teaching of being taught in schools. The resident disagrees (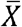= 2.13) with the position that issues relating to crime, unemployment and cost of living are more important than keeping a garbage-free community. The residents agreed further (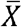=2.04) to the idea of picking up waste around their community as one of the ways by which responsibility to the nation.

However, the residents demonstrated negative attitudes (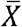= 1.89) in terms of indiscriminate dumping of waste on the streets and in the drains because they did not have waste receptacles to dispose it. Lastly, they did not see the responsibility of the local government authority in waste collection and management (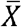= 1.59). From the results presented above, it is clear that the residents of Ijebu-Ode community were favourably disposed to collection and disposal of solid waste in their neighbourhood. Inspite of this, pockets of dirty locations and sites were still found, perhaps there were constraints that negate their positive disposition to waste management, these may likely be poor economic empowerment, poor level of awareness of the effective waste management procedure and lack or inadequate waste disposal agencies. The result is similar to that of (Bowersox et. al., 2005; Johansson, 2006; Thrift, 2007) argues that improper/negative attitude of individual towards waste management may influence lack or inadequate awareness of individual, inadequate waste collection method etc. and states that for a positive attitude of people towards waste management, proper awareness of must be built among the communities supports with increase in level education measures, improved waste collection service and availability of fund. By this result, Browne and Allen (2007) is considered to have in relation to the aspect of their work that addresses positive influence of attitude towards solid waste disposal through awareness-building campaigns and educational measure on the negative impacts of inadequate waste collection with regard to public health and environmental conditions, and the value of effective disposal.

**Figure 3.**
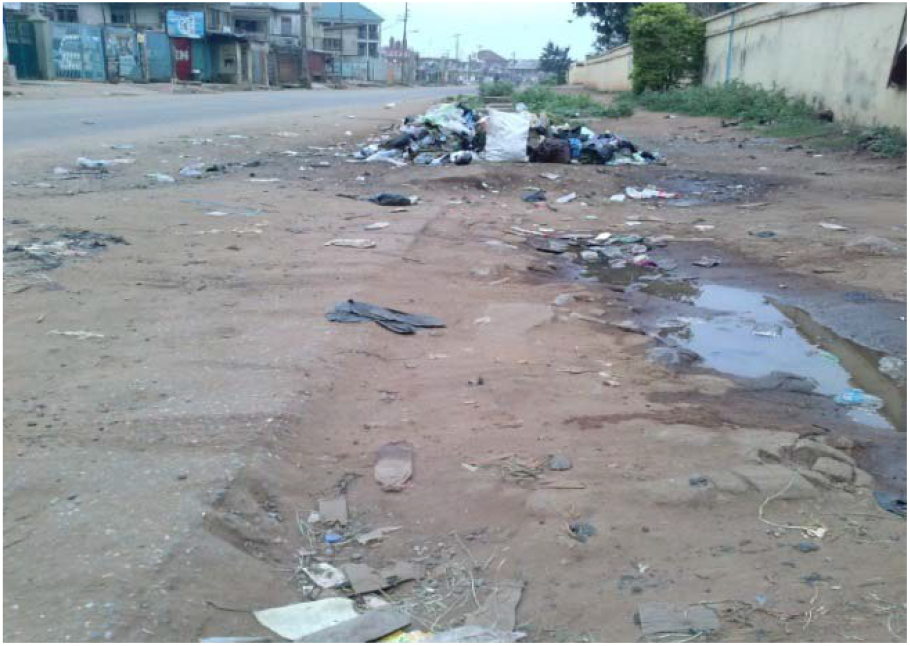
Study area Source: Field capture, 2018.

**Figure 4.**
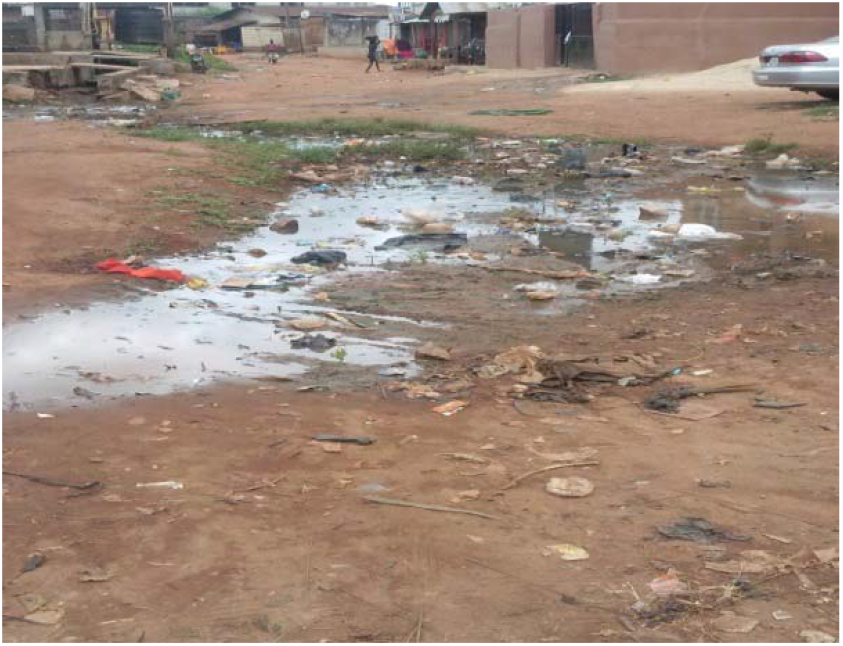
Study area Source: Field capture, 2018.

### Result on how waste being disposed by residents of Ijebu-Ode metropolis?

The results on table 2 shows that the residents of residents of Ijebu-Ode metropolis generally employed the use of single bin in collection of their wastes.

**Table 2.** method of collection of domestic waste

| S/N | Item | Median | S.D | Decision |
| --- | --- | --- | --- | --- |
| 1 | What method do you use for storage of domestic waste at your home? | 2.00 | .67 | Single bin |

(Median =2.00)

Given the result (median = 2.00) on table 3, wastes from homes of the residents of Ijebu-ode community were disposed on a generally note twice a month.

**Table 3.** Frequency of collection of waste in a month

| S/N | Item | Median | S.D | Decision |
| --- | --- | --- | --- | --- |
| 1 | How many times waste is collected from your home in a day? | 2.00 | 1.08 | Twice |

Looking at the results (Median= 1.00) on table 4, the waste disposal procedure among the residents of Ijebu-Ode metropolis was the use of common waste receptacles placed in designated locations in the community.

**Table 4.** Disposal approach of domestic wastes

| S/N | Item | Median | S.D | Decision |
| --- | --- | --- | --- | --- |
| 1 | Which collection system is adopted in your locality to collect the solid waste? | 1.00 | .50 | Common receptacles in designated locations |

The use of single bin in collection of waste appears inadequate coupled with twice frequency of disposal of wastes in a month. The implication is that the tendency of waste spilling over the bin to litter the surrounding before being disposed is high. The use of common receptacles at designated spots in the community is also not too appropriate, because the health of people closed to these spots stands the risk of health challenge. As it also noted from the results above, wastes from homes of the residents of Ijebu-Ode community were disposed on a generally note twice a month, other respondents also used several unlawful methods to get rid of the waste like dumping into gutter, burning, dumping on undeveloped land, while few others buried theirs. These confirm the findings of Franduah’s (2008) that majority of the population dumps their refuse in unauthorized sites. This could be due to inadequacy of services provided by the waste managers. Also, this correlate with that of Banjo et. la., (2009) that states that majority of the residents of Ijebu disposed their waste into the environment, while few of has single bin of keeping waste

### Adequate of health status of Ijebu-Ode residents

Looking at the results (Median= 1.00) on table 5 residents of Ijebu-Ode community claimed awareness of health related diseases of waste disposal.

**Table 5.** Awareness of health-related diseases of waste disposal

| S/N | Item | Median | S.D | Decision |
| --- | --- | --- | --- | --- |
| 1 | Are you aware of health-related disease of waste disposal before? | 1.00 | .34 | Aware |

The result (Median = 1.00) on table 6 show that the frequency of outbreak of diseases as a result of keeping refuse dumps with residential areas was low.

**Table 6.** Frequency of outbreak of diseases due to refuse dump sites in the community

| S/N | Item | Median | S.D | Decision |
| --- | --- | --- | --- | --- |
| 1 | What is the frequency of outbreak of diseases as a result of keeping dumping sites in your community? | 1.00 | .50 | Low |

From the results (Median = 0.00) on table 7, residents of Ijebu ode community had experience of infectious disease at one time or the other in the past.

**Table 7.** Residents’ experience of infectious diseases

| S/N | Item | Median | S.D | Decision |
| --- | --- | --- | --- | --- |
| 1 | Have you experience any infectious disease before? | 0.00 | .50 | Positive |

The result (Median = 1.00) on table 8 on the frequency of treatment received by the residents of Ijebu ode of the same infectious disease was low.

**Table 8.** Frequency of some infectious disease treatment by the residents.

| S/N | Item | Median | S.D | Decision |
| --- | --- | --- | --- | --- |
| 1 | How many time you have treated this same disease? | 1.00 | .47 | Low |

According to the result (Median = 0.00) on Table 9, the awareness of the residents of Ijebu-Ode community of other residents infectious disease was positive.

**Table 9.** Awareness of other residents with experience of infectious disease

| S/N | Item | Median | S.D | Decision |
| --- | --- | --- | --- | --- |
| 1 | Do you know someone in your area that have experience same disease or nay infectious disease? | 0.00 | .51 | Positive |

Considering the results (Median = 0.00) on Table 10 on suggestion about what to do to prevent occurrence of infectious diseases was nil concerning the residents of Ijebu-ode community.

**Table 10.** Awareness of residents of Ijebu-ode on the infectious diseases

| S/N | Item | Median | S.D | Decision |
| --- | --- | --- | --- | --- |
| 1 | Is there any suggestion given to you concerning this disease? | 0.00 | .49 | None |

From the above results, it obvious that the residents of Ijebu-Ode are aware of health related diseases such as typhoid, diarrhea, fever etc. and the frequency of outbreak of these diseases in the community is low, but majority of the residents have experienced these health related disease with low frequency, that is, many experienced these diseases either once or twice but no frequent or continuous nursing of either same disease or another. Also, the residents are aware of neighbour or other residents that experienced any of the diseases before. Which may be conclude that the residents have experienced health related diseases before either as a result of keeping of dirty environment and other factors that may account. However, this results support the argument of Mosquera et. al. (2009) that people in developing countries hold the perception that improper solid waste disposal results in negative health outcomes yet. Also this result support the similar work done in the same study area (Ijebu-Ode) by Banjo et. al., (2009) stated that majority of the residents have experienced different infectious disease as result of keeping dumping site around the houses and environment at large.

### TESTING HYPOTHESIS

**Ho**_**1**_: There is no significant relationship between waste management mechanism and health status of residents.

Given the result of test of relationship (Spearman rho= 0.92; p (.318)> .05) on table 11, there is no significant relationship between waste management mechanism and health status of Ijebu-ode residents. Thus, the null hypothesis is not rejected. Thus, the residents have experienced many health related diseases but might not directly influence by indiscriminate dumping or keeping dirty environment. Other factors such natural or other human activities that is hazardous to human health may account for this. Although, the waste management system in the area is low, these do not have high implication on their health, but the result shows that their healthiness or well being do not really depend on waste management method. This result opposes the findings of an earlier study involving the whole Ijebu-Ode conducted by Banjo et. al. (2009) which states that keeping of dumping site or waste management system in the area have a great implication on the health of the resident of the community.

**Table 11.**
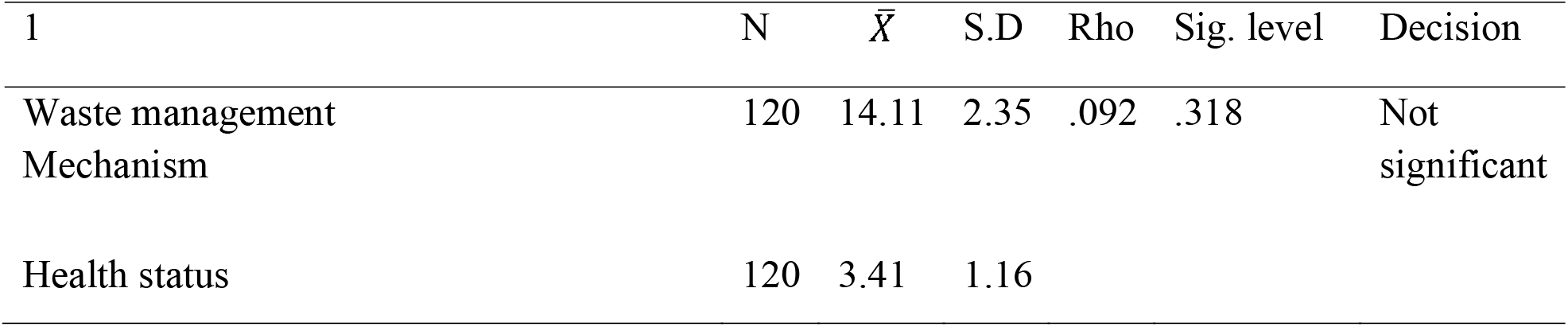
Spearman rho indicates relationship between waste management mechanism and health status of Ijebu-ode residents

| 1 | N | $\bar{X}$ | S.D | Rho | Sig. level | Decision |
| --- | --- | --- | --- | --- | --- | --- |
| Waste management Mechanism | 120 | 14.11 | 2.35 | .092 | .318 | Not significant |
| Health status | 120 | 3.41 | 1.16 |  |  |  |

Further, the result obtained indicate that indiscriminate dumping or poor waste management is not majorly determinant of being healthy while some other factors such cultural activities, economic activities, accident, water consumption etc. may account to health status of an individual. This opposes the finding of Abul (2010) which states the low health status or poor health majorly determine by dirty environment by poor waste management.

The reason for this result could be traced to an interplay of factors of involving improved human health and good living condition with safe and clean environment. Periodic and consistent monitoring and management waste by Ogun State Environmental Protection Agency (OGEPA) and Local authorities with continuous education and awareness of individual living in the community and the state at large on better waste management mechanism.

## CONCLUSION AND IMPLICATION OF STUDY

The study empirically examined the waste management mechanism as determinant of health status of the residents of Ijebu-Ode metropolis. From the findings all the objectives were accomplished. For the attitude of respondents towards waste management, with the aggregate mean (2.29) show that the respondents expressed that there is positive attitude of residents towards waste management, and also prove that they are favourably disposed to collection and disposal of solid waste in their neighbourhood. Inspite of these their environment is still found to be littered with indiscriminate waste across the streets. However, the knowledge of the residents maybe regard as general / residual knowledge on environment, not that they fully have the understanding of waste disposal and management.

It is however concluded that there are many factors that may attribute to health problem among individual except of improper or poor waste management. The outcome of this research gives an insight that health problem in any area; there will be other factor that is accountable for it. This affirmation is given as regard of this research which the results shows that there is no significant relationship between the waste management and health status. Not that health may not affect by poor waste management, but not in totality.

On general conclusion, it is obvious that the residents do not have full understand or knowledge of waste management but rather base on residual knowledge of waste management and other little experience gotten from their immediate environment. That is why pile of waste is found across streets of the community. Thus, the effort of the local authorities and state government and private stakeholders towards managing the environment is not sufficient. Stakeholders need to improve their efficient on the managing good and hygienic environment for sustainability. Findings from this study have implications for stakeholders as follow:

i. The federal and state ministries with private and local authorities of Environment should be proactive to increase the waste management mechanism and monitoring the environment because if it continue this way, there will be more threat on environment
ii. Concerted efforts of the government, policy makers and individuals towards waste disposal and management is very much important for a sustainable development and improve and continuous clean, safe and healthy environment and inhabitants.

## RECOMMENDATION

The following recommendations are suggested based on the study as follows;

i. The State and Local Authorities should provide a number of waste bins at vantage areas in the various communities. They should be supplied with enough containers to avoid indiscriminate dumping of waste in gutters, open spaces, streets and nearby bushes.
ii. There should be public education on proper ways of solid waste disposal in the metropolis to inform the general public on the implications of unhealthy environment and the need to keep their communities clean. The education could be done by Local Authorities and the Ogun State Environmental Programme Agency (OGEPA).
iii. The environmental management authority should make it a responsibility of introducing the use of standard bins with lid at all levels for domestic and commercial use to the people most especially along the street so as to reduce the probability of people littering the environment.

## DECLARATIONS

### Ethics approval

Ethics Committee of Tai Solarin University of Education, Ijagun, Ogun State, Nigeria gave ethical approval for this work.

### Consent to participate

Written informed consent was obtained from all participants prior to data collection.

### Funding

This research did not receive any specific grant from funding agencies in the public, commercial, or not-for-profit sectors.

### Competing interests

The authors declare that they have no competing interests.

## Data availability

The data supporting the findings of this study are available from the corresponding author upon reasonable request.

## Notes

### Competing Interest Statement

The authors have declared no competing interest.

### Funding Statement

This study did not receive any funding

